# Exposure to SARS-CoV-2 within the household is associated with greater symptom severity and stronger antibody responses in a community-based sample of seropositive adults

**DOI:** 10.1101/2021.03.11.21253421

**Authors:** Joshua M. Schrock, Daniel T. Ryan, Rana Saber, Nanette Benbow, Lauren A. Vaught, Nina Reiser, Matthew P. Velez, Ryan Hsieh, Michael Newcomb, Alexis R. Demonbreun, Brian Mustanski, Elizabeth M. McNally, Richard D’Aquila, Thomas W. McDade

## Abstract

Magnitude of SARS-CoV-2 virus exposure may contribute to symptom severity. In a sample of seropositive adults (n=1101), we found that individuals who lived with a known COVID-19 case exhibited greater symptom severity and IgG concentrations compared to individuals who were seropositive but did not live with a known case (P<0.0001).

## Introduction

SARS-CoV-2 infections are highly variable in both disease severity and magnitude of antibody response following infection.^1,2^ It has been hypothesized that the magnitude of SARS-CoV-2 viral exposure (amount of virus inoculum, closer contact, more prolonged exposure) may contribute to disease severity.^3-5^ This relation between exposure dose and disease severity has been demonstrated experimentally for other respiratory viruses, including influenza.^6^ In addition, higher SARS-CoV-2 viral load during acute infection in humans has been linked to greater disease severity.^7^

Greater disease severity, in turn, may be associated with stronger antibody responses in the convalescent phase of infection.^8-10^ We previously reported that those who reported more symptoms had higher concentrations of immunoglobulin G (IgG) antibodies in a community-based sample of seropositive adults.^10^ Another study using clinical assessments of disease severity reported similar findings.^9^

Respiratory virus exposures within the household may often be more intense than respiratory virus exposures outside of the household.^3,5^ For example, studies of measles epidemics have reported that cases acquired from a cohabitant have higher age-specific case fatality ratios compared to cases acquired outside the home.^11,12^ Exposure to SARS-CoV-2 within the household may be more prolonged, physically proximate, and unmitigated by personal protective equipment compared to transient public exposures.^3,5^ While some may attempt to isolate from cohabitants when they develop symptoms of COVID-19, it has been demonstrated that high levels of SARS-CoV-2 viral shedding occur prior to the onset of symptoms.^13^ Furthermore, isolation within the household may not be achievable in many living situations.

The aim of this paper is to compare symptom severity and SARS-CoV-2 IgG antibody concentrations in seropositive individuals who shared living space with a known COVID-19 case vs. individuals who are seropositive but did not share living space with a known COVID-19 case.

## Methods

The analytic sample in this report (n=1101) consists of participants who screened positive for SARS-CoV-2 in the Screening for Coronavirus Antibodies in Neighborhoods (SCAN) study.^2,10,14,15^ SCAN has administered online surveys and collected dried blood spot (DBS) samples from >5000 participants. Recruitment messages were disseminated through social media, email blasts, print flyers, newspaper advertisements, and local press coverage. Participants were recruited from neighborhoods throughout the Chicago area and from personnel of the Northwestern University Feinberg School of Medicine (FSM) in Chicago. Eligible participants who consented to participate completed an online survey and received a kit for self-collection of a DBS sample. DBS kits were either sent to participants through the mail or, for FSM participants, made available for pickup. All study protocols were approved by the IRB at Northwestern University (#STU00212457 and #STU00212472).

IgG antibodies to the receptor binding domain of SARS-CoV-2 were quantified using an enzyme-linked immunosorbent assay (ELISA) that has received emergency use authorization from the FDA.^16,17^ We adapted and validated this assay for use with DBS samples.^14^ The cut-off for seropositivity was set at the optical density value for to the 0.39 μg/ml calibrator.^14^

Participants were presented a checklist of symptoms and asked to report whether they had experienced each symptom since March 1, 2020. In a previous study, we identified a cluster of eight symptoms that were associated with higher SARS-CoV-2 IgG concentrations.^10^ To create a symptom severity score, we weighted each of the eight symptoms by its regression coefficient in a simple regression model with the symptom as the independent variable (1=present, 0=absent) and log_2_ IgG concentration as the dependent variable. In other words, symptoms more strongly associated with IgG levels were assigned larger weights. Symptoms were assigned the following weights: loss of taste/smell=1.05, fever=0.69, muscle/body aches=0.61, shortness of breath=0.49, fatigue/excessive sleepiness=0.46, diarrhea/nausea/vomiting=0.43, cough=0.41, and headache=0.26. The resulting symptom severity score ranged from 0 to 4.40 (mean=1.10, SD=1.22). Similar weighting schemes have been used in prior research to generate quantitative symptom severity scores.^18,19^

Exposure to cohabitants with COVID-19 was assessed by asking the following question, “Since March 1, 2020, has anyone in your household been told by a healthcare provider that they have, or likely have, COVID-19? Do not include yourself when answering this question.”

The covariates included in statistical models were age, sex assigned at birth, racial/ethnic identity, chronic pre-existing conditions (having one or more of the following: chronic kidney disease, chronic lung disease, diabetes mellitus, cardiovascular disease, or body mass index >30 kg/m2; 1=yes, 0=no), tobacco use (since March 1^st^, 2020; 1=yes, 0=no), working outside the home in close proximity to others (since March 1^st^, 2020; 1=yes, 0=no), number of cohabitants in the household, and date of inclusion in the study (number of days since March 1^st^ that the DBS kit was received at the lab).

We fitted ordinary least squares regression models with exposure to cohabitants with COVID-19 as the independent variable, with the covariates described above, and with symptom severity scores and log_2_ IgG concentrations as the dependent variables. These models were fitted using “lm” function in base R (version 4.0.3).

## Results

The median date of inclusion in the study was October 23^rd^, 2020 (Range: June 30^th^, 2020 to January 20^th^, 2021). None of the study participants reported having been vaccinated for SARS-Cov-2 at the time of survey completion. Out of 1101 seropositive participants, 42.7% identified as non-Hispanic white, 23.9% as Hispanic/Latinx, 8.5% as non-Hispanic Black, 18.9% as Asian, and 6% selected other response categories for race/ethnicity. The mean age was 38.62 (SD=12.55), and 56.2% were assigned female at birth. Symptom severity scores were higher among participants who had been exposed to a cohabitant diagnosed with COVID-19 (mean=2.54, SD=1.46) compared to participants who not been exposed to a cohabitant diagnosed COVID-19 (mean=0.93, SD=1.08) (t=25.83, P<0.0001). Concentrations of SARS-CoV-2 IgG were also higher among participants who had been exposed to a cohabitant diagnosed with COVID-19 (mean=2.21 ug/mL, SD=2.27) compared to participants who not been exposed to a cohabitant diagnosed with COVID-19 (mean=1.25 ug/mL, SD=1.54) (t=24.34, P<0.0001) **(Figure 1)**.

**Figure 1.**
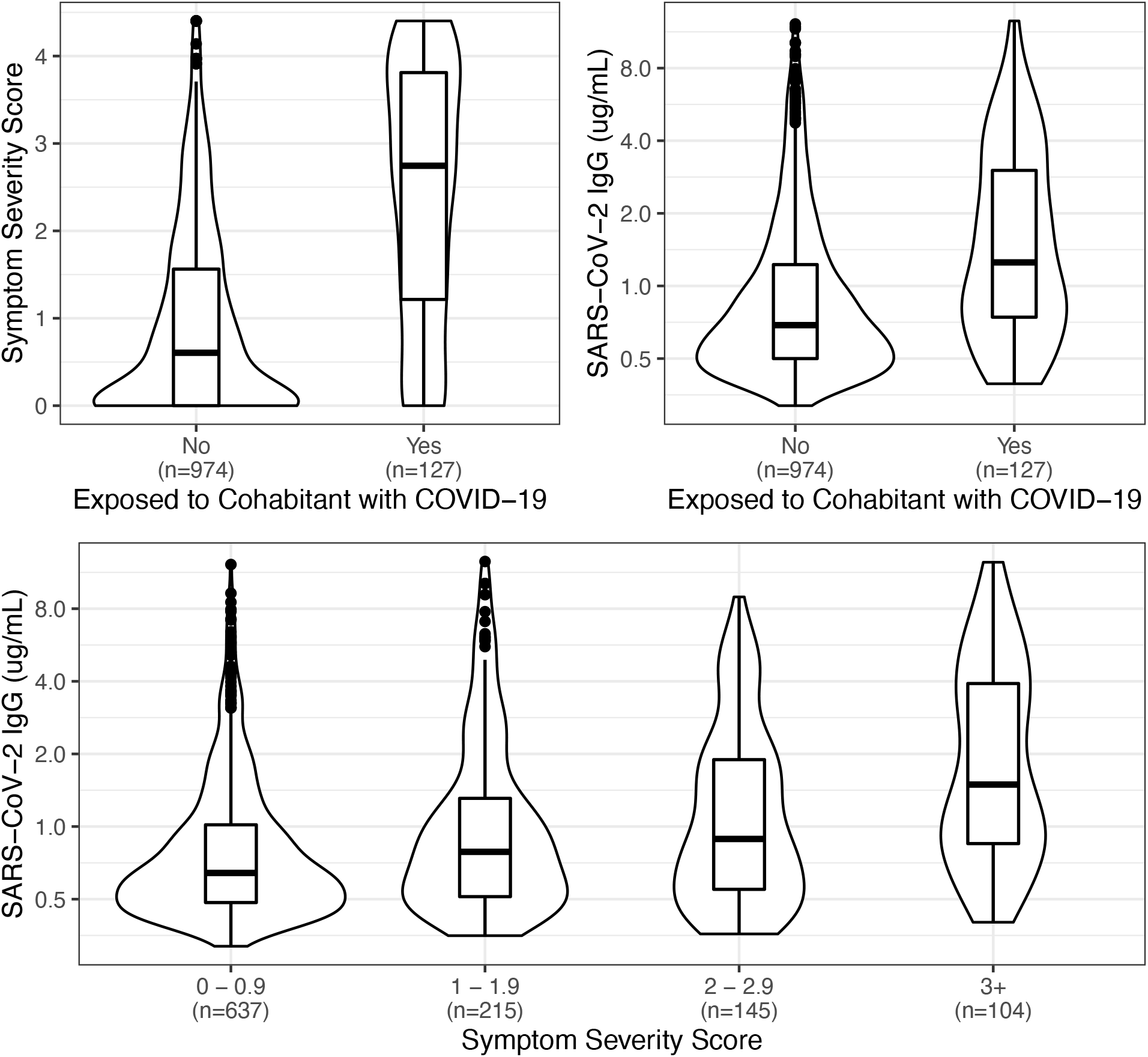
Associations between exposure to a cohabitant with Covid-19, symptom severity scores, and SARS-CoV-2 antibody concentrations. The y-axis for SARS-CoV-2 IgG concentrations is presented on a log_2_ scale. Data are from a community-based sample of seropositive adults from the Chicago area (n=1011). The distribution for each category is represented by smoothed kernel density plots overlaid with boxplots depicting interquartile ranges. This figure was created using the R package “ggplot2” (version 3.3.2).

In a regression model adjusted for age, sex, race/ethnicity, chronic pre-existing conditions, tobacco use, working in proximity to others, number of cohabitants, and date of inclusion in the study, we found that having been exposed to a cohabitant diagnosed with COVID-19 was associated with higher symptom severity scores (β=1.35, SE=0.106, P<0.0001). In a second regression model adjusted for the same covariates, we found that having been exposed to a cohabitant diagnosed with COVID-19 was associated with higher concentrations of log_2_ IgG (β=0.7, SE=0.105, P<0.0001). Unstandardized coefficients and standard errors for both models are presented in **Table 1**.

**Table 1.** Coefficients, with standard errors in parentheses, from ordinary least squares regression models with symptom scores and log_2_ SARS-CoV-2 IgG levels as outcome variables. Data are from sample of adults seropositive for SARS-CoV-2 in the Chicago area (n=1101). This table was created using the R package “stargazer” (version 5.2.2).

|  | *Outcome variable:* |  |
| --- | --- | --- |
|  | Symptom Severity Score | Log<sub>2</sub> IgG Concentration (ug/mL) |
| Age | -0.002<br>(0.003) | **0.007**<br>**(0.003)** |
| Assigned female at birth | 0.088<br>(0.069) | 0.122<br>(0.068) |
| Race/ethnicity (ref=white) |  |  |
| Hispanic/Latinx | **0.272**<br>**(0.087)** | **0.186**<br>**(0.086)** |
| Black | 0.180<br>(0.126) | **0.249**<br>**(0.124)** |
| Asian | -0.168<br>(0.093) | -0.159<br>(0.092) |
| Other | 0.265<br>(0.145) | 0.256<br>(0.143) |
| Chronic pre-existing condition | **0.370**<br>**(0.078)** | 0.039<br>(0.077) |
| Tobacco use | 0.219<br>(0.134) | 0.023<br>(0.132) |
| Working in proximity to others | -0.094<br>(0.069) | 0.013<br>(0.068) |
| Number of cohabitants | 0.002<br>(0.024) | **-0.046**<br>**(0.023)** |
| Date of inclusion in the study | **0.002**<br>**(0.001)** | **0.002**<br>**(0.001)** |
| Cohabitant with COVID-19 | **1.353**<br>**(0.106)** | **0.704**<br>**(0.105)** |
| Intercept | 0.395<br>(0.209) | **-1.008**<br>**(0.206)** |
| Observations | 1,101 | 1,101 |
| R<sup>2</sup> | 0.208 | 0.084 |

*Note:* Coefficients statistically significant at P<0.05 are in bold

## Discussion

In models adjusted for key covariates, we found that seropositive individuals who reported having a cohabitant with COVID-19 had substantially greater symptom severity and IgG antibody responses compared to seropositive individuals who did not report having a cohabitant with COVID-19. Notably, working in close proximity to others, which has received substantial attention as a risk factor for COVID-19,^20^ did not exhibit statistically significant associations with symptom severity or IgG antibody concentrations. Our results suggest that high virus exposure levels, which may often occur when exposed to a cohabitant with COVID-19, contribute to greater disease severity and stronger antibody responses.

It is worth noting that our study design does not allow us to determine whether the cohabitant diagnosed with COVID-19 was the first instance of SARS-CoV-2 exposure within the household. In some cases, the study participant may have been exposed prior to the cohabitant. Future studies should include immunological data on multiple household members and track the sequence of seroconversion and symptom occurrence.

Regardless of which individual in the household was exposed first, our results suggest that preventing transmission within households should be a critical area of focus for public health efforts designed to reduce rates of symptomatic COVID-19. Policies and interventions that apply only to public places (e.g., mask mandates, business capacity limits) may be insufficient unless they are combined with measures that also reduce transmission within households (e.g., intensive testing, contact tracing, and isolation programs).

It is also important to consider the implications of exposure dose when evaluating possible changes in vaccination strategies. Recent preprints have reported strong immune responses following a single dose of an mRNA vaccine among individuals who were seropositive prior to infection.^21,22^ Based on these findings, some have suggested forgoing the second vaccine dose for those who have prior exposure.^21,22^ However, the participants in these studies were individuals who qualified for early vaccination. One of the reports only studied health care workers (HCW).^22^ In that report, HCW with higher pre-vaccine antibody levels had a greater magnitude of post-vaccine antibody response.^22^ The other report did not report pre-vaccine antibody levels and did not describe the study sample in detail.^21^ The priority 1a group, which was being vaccinated at the time the study took place, consisted of frontline HCWs and/or long-term care facility residents, which are each at high risk for close, prolonged, and repeated virus exposure.^23^ Robust antibody responses following a single vaccine dose may or may not generalize to seropositive individuals who had milder disease or lower levels of pre-vaccination antibodies resulting from prior natural infection. The results of our study demonstrate the importance of broad sampling strategies that capture an adequate range of variation in exposure context, symptom severity, and acquired immunity.

## Data Availability

Data are available upon request from the corresponding author subject to an approved data sharing agreement to protect participant confidentiality.

## Acknowledgements

This material is based upon work supported by the National Science Foundation under Grant No. 2035114 to TWM, RD, EMM, BM, and ARD. Additional support was provided by the Northwestern University Office of Research, the Northwestern University Clinical and Translational Sciences Institute (NIH UL1TR001422), and a generous gift from Dr. Andrew Senyei and Noni Senyei. The funding sources had no role in the study design, data collection, analysis, interpretation, or writing of the report.

